# Dynamic Lymphocyte Recovery Patterns Predict 90-Day Mortality in Sepsis: A Machine Learning-Enhanced Analysis of the MIMIC-IV Cohort

**DOI:** 10.1101/2025.11.11.25339967

**Authors:** Yuwei Huang, Yuzheng Zhang, Zhilei Fan, Qiong Chen, Yongli Gao

## Abstract

**Background:** Sepsis-induced immunosuppression, characterized by lymphopenia, is associated with adverse outcomes. We aimed to identify distinct lymphocyte recovery patterns in patients with sepsis, evaluate their association with mortality, and develop a machine learning model to enhance prediction

**Methods:** This retrospective cohort study included adult patients with sepsis and initial lymphopenia (Absolute Lymphocyte Count [ALC] < 1.0 × 10L/L) from the Medical Information Mart for Intensive Care (MIMIC)-IV database. We defined three lymphocyte recovery patterns: "Persistent Suppression," "Partial Recovery," and "Complete Recovery." The primary outcome was 90-day all-cause mortality. Multivariable Cox models were used to assess the association between recovery patterns and mortality. An Extreme Gradient Boosting (XGBoost) model was developed to predict 90-day mortality.

**Results:** 90-day mortality was highest in the Persistent Suppression group (49.1%) versus Partial (41.3%) and Complete Recovery (35.7%) groups (p<0.001). Persistent Suppression remained an independent predictor of mortality (adjusted Hazard Ratio 1.31, 95% CI 1.10-1.55). The XGBoost model achieved superior discrimination (Area Under the Receiver Operating Characteristic Curve [AUROC]=0.767) over traditional scores. SHAP analysis confirmed that dynamic features, including lymphocyte recovery metrics, were key model drivers. The model also demonstrated robust performance across various clinical subgroups (e.g., age, disease severity).

**Conclusions:** The trajectory of lymphocyte recovery following sepsis onset is an independent prognostic marker. Failure to restore lymphocyte counts is strongly associated with increased long-term mortality. Integrating this dynamic immunological feature into machine learning algorithms significantly enhances predictive accuracy, offering a promising tool for real-time risk stratification.

## Introduction

Sepsis is a life-threatening organ dysfunction caused by a dysregulated host response to infection and represents a major cause of mortality and critical illness worldwide[1, 2]. Despite advances in supportive care, the incidence of sepsis continues to rise, and it remains a leading driver of healthcare costs[3].

The pathophysiology of sepsis is complex, involving a biphasic immune response that often transitions from an initial hyper-inflammatory "cytokine storm" to a protracted state of immunosuppression[4, 5]. This latter phase, termed sepsis-induced immunosuppression, is characterized by the depletion and functional exhaustion of immune effector cells, particularly lymphocytes[6]. Profound and sustained lymphopenia is a hallmark of this condition and has been consistently associated with an increased risk of secondary infections, prolonged organ dysfunction, and higher mortality[7].

Current clinical practice relies on severity-of-illness scores, such as the Sequential [Sepsis-related] Organ Failure Assessment (SOFA) score, and biomarkers like lactate to stratify risk and guide therapy[1]. While useful, these tools primarily reflect the degree of organ dysfunction and tissue hypoperfusion, offering only a static snapshot of the patient’s condition at a specific moment. Simple hematological indices, such as the neutrophil-to-lymphocyte ratio (NLR), have also emerged as valuable prognostic markers, highlighting the importance of the host’s immune status[8, 9]. However, these static measurements fail to capture the dynamic evolution of the host’s immune response over the course of the illness.

We hypothesized that the trajectory of lymphocyte count recovery following the initial septic insult holds more significant prognostic information than a single baseline measurement. The ability or failure to restore lymphocyte populations may reflect the underlying capacity of the host’s immune system to recover from the septic challenge. Therefore, this study aimed to: 1) define distinct patterns of lymphocyte recovery in a large, contemporary cohort of critically ill patients with sepsis using the MIMIC-IV database; 2) determine the association of these recovery patterns with 90-day mortality; and 3) develop and validate a machine learning model incorporating these dynamic features to improve prognostic accuracy.

## Methods

### Study Design and Data Source

We conducted a retrospective cohort study using data from the Medical Information Mart for Intensive Care (MIMIC)-IV database (version 3.1), a large, publicly available, de-identified database containing comprehensive clinical data of patients admitted to intensive care units at the Beth Israel Deaconess Medical Center in Boston, MA[10]. The study was conducted in accordance with the STROBE (Strengthening the Reporting of Observational Studies in Epidemiology) guidelines.The use of this database was approved by the institutional review boards of the Massachusetts Institute of Technology and Beth Israel Deaconess Medical Center. The authors obtained access to the MIMIC-IV database after completing the CITI ’Data or Specimens Only Research’ course (Record ID: 66998740) and signing a data use agreement (DUA) with PhysioNet.

### Study Population

We included adult patients (age ≥ 18 years) on their first ICU admission who met the Sepsis-3 criteria, defined as a suspected infection with a concurrent acute increase in the Sequential [Sepsis-related] Organ Failure Assessment (SOFA) score of 2 points or more[1]. We further selected patients who developed lymphopenia (Absolute Lymphocyte Count [ALC] < 1.0 x 10L/L) within the first 72 hours of ICU admission, a commonly used threshold to define lymphopenia in sepsis studies[11]. To ensure adequate observation of the lymphocyte trajectory, we required an ICU length of stay greater than 72 hours and at least three ALC measurements within the first 7 days of their ICU stay. Patients were excluded if they had a diagnosis of hematological malignancy or HIV, were on long-term immunosuppressive or chemotherapy agents prior to admission, or had critical data missing for exposure or outcome assessment. These exclusion criteria are standard in sepsis cohort studies using electronic health records to minimize confounding from pre-existing profound immunosuppressive conditions[12]. These criteria, necessary for tracking the lymphocyte trajectory, inherently introduce a potential for survivorship bias by excluding patients who died or were discharged within the first 72 hours, a limitation addressed in the discussion.

### Variables and Definitions

The primary exposure was the **lymphocyte recovery pattern**, defined based on ALC trajectory from its nadir to day 7 of ICU admission. The thresholds for defining recovery patterns were informed by previous studies that have identified ALC levels around 0.8 x 10L/L and 1.2 x 10L/L as clinically relevant cutoffs for severe lymphopenia and the lower limit of the normal range, respectively, in critically ill populations[13]. Patients were categorized into three mutually exclusive groups:

1. **Complete Recovery (Reference Group):** ALC recovered to ≥ 1.2 x 10L/L at least once.
2. **Partial Recovery:** ALC recovered to ≥ 0.8 x 10L/L but never reached 1.2 x 10L/L.
3. **Persistent Suppression:** ALC never recovered to ≥ 0.8 x 10L/L.

The primary outcome was **90-day all-cause mortality**. Secondary outcomes included 28-day ventilator-free days (VFDs) and ICU length of stay (LOS). Covariates for adjustment were selected based on clinical relevance and included demographics, comorbidities (Charlson Comorbidity Index), and markers of initial illness severity.

### Dynamic Feature Engineering

To capture the temporal changes of key biomarkers for the predictive model, several dynamic variables were engineered from time-series data within the first 7 days of ICU admission. For instance, "Platelet Slope" was calculated as the slope of the linear regression line fitted to all platelet count measurements during this period. Similarly, "ALC Recovery Ratio" was defined as the ratio of the maximum ALC to the minimum ALC within the first 7 days. These derived features were designed to represent the trajectory and magnitude of change in a patient’s physiological state.

### Statistical Analysis

A two-stage analysis was performed using Python (version 3.10).

### Stage 1: Association Analysis

Baseline characteristics were compared across the three recovery pattern groups. To assess the association between lymphocyte recovery patterns and 90-day mortality, we constructed a series of multivariable Cox proportional hazards models[14]. For secondary outcomes, we used Fine-Gray competing risk regression to analyze the association with ICU LOS, treating death as a competing event, which is essential for obtaining an unbiased estimate when a competing event is present[15].

### Stage 2: Predictive Modeling

The dataset was randomly split into a training set (70%) and a testing set (30%). We developed an Extreme Gradient Boosting (XGBoost) model, a powerful and widely used machine learning algorithm for clinical prediction tasks[16]. The model’s performance was evaluated on the testing set using the Area Under the Receiver Operating Characteristic curve (AUROC). Model interpretability was assessed using SHapley Additive exPlanations (SHAP), a state-of-the-art method to explain the output of any machine learning model by attributing the prediction to each feature[17].

## Results

### Patient Characteristics

The patient selection process is detailed in the study flowchart (Figure 1). From an initial screening of patients meeting Sepsis-3 criteria, a final cohort of 1,965 patients who developed lymphopenia within 72 hours of ICU admission was included for analysis. Based on their lymphocyte recovery trajectories, patients were stratified into three groups: Persistent Suppression (n=576, 29.3%), Partial Recovery (n=554, 28.2%), and Complete Recovery (n=835, 42.5%).

**Figure 1.**
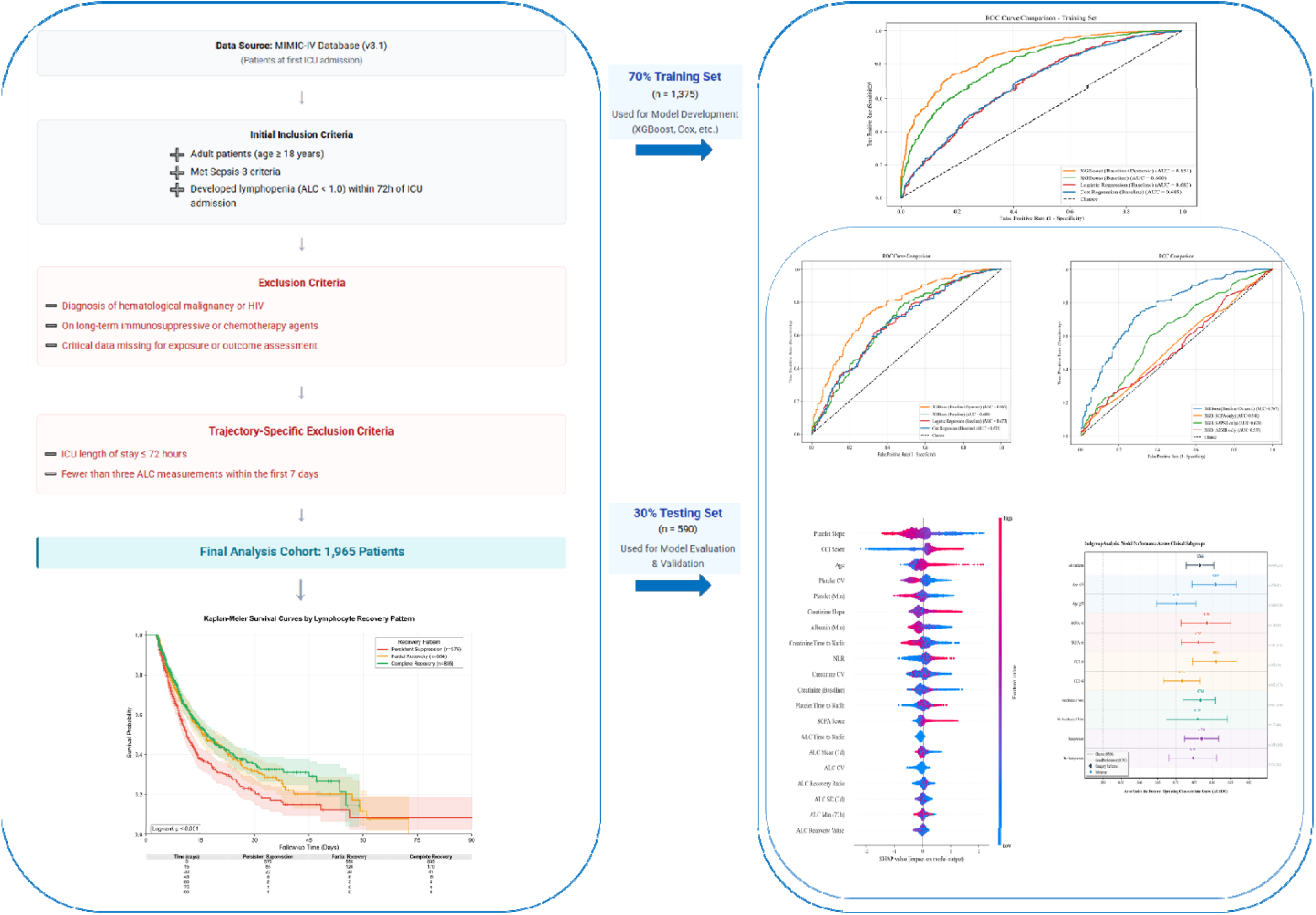
Flowchart of study design. This flowchart illustrates the patient inclusion and exclusion criteria. Starting from patients admitted to the ICU in the MIMIC-IV database, the cohort was filtered based on Sepsis-3 criteria, age, initial lymphopenia (ALC < 1.0 x 10LJ/L within 72h), ICU stay duration, and availability of ALC measurements, resulting in the final cohort for analysis.

The baseline demographic and clinical characteristics of the study population are summarized in Table 1. Significant differences were observed among the three groups. Compared to the Complete Recovery group, patients in the Persistent Suppression group were significantly older (mean age, 67.6 vs. 62.0 years), had a greater comorbidity burden as indicated by a higher Charlson Comorbidity Index (6.3 vs. 4.8), and presented with a higher severity of illness at admission, reflected by a higher SAPS II score (46.8 vs. 44.9) (all p < 0.05). Furthermore, patients with persistent suppression exhibited a significantly lower baseline platelet count (137.2 vs. 173.8 ×10L/L, p < 0.001).

**Table 1.**
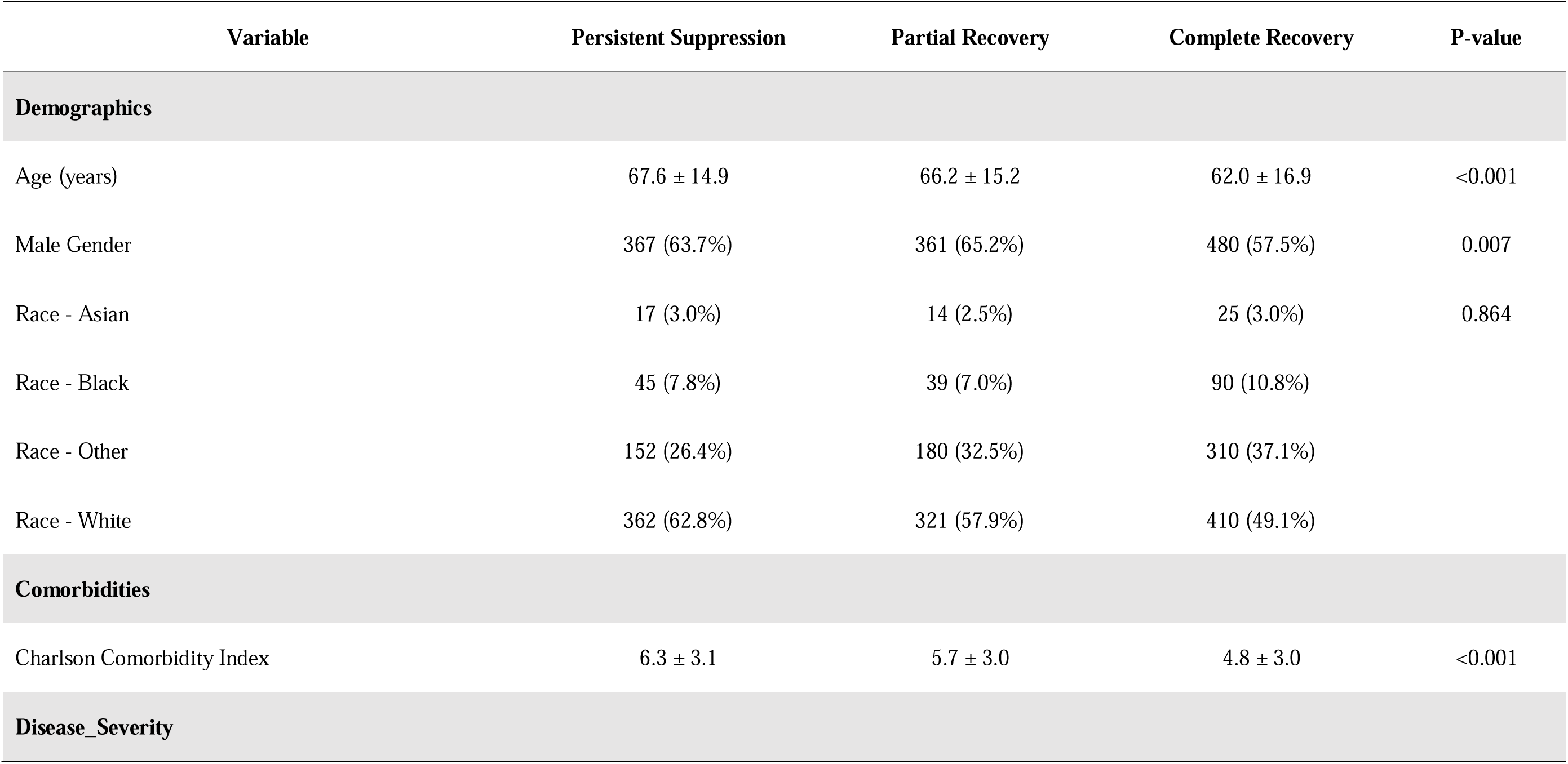
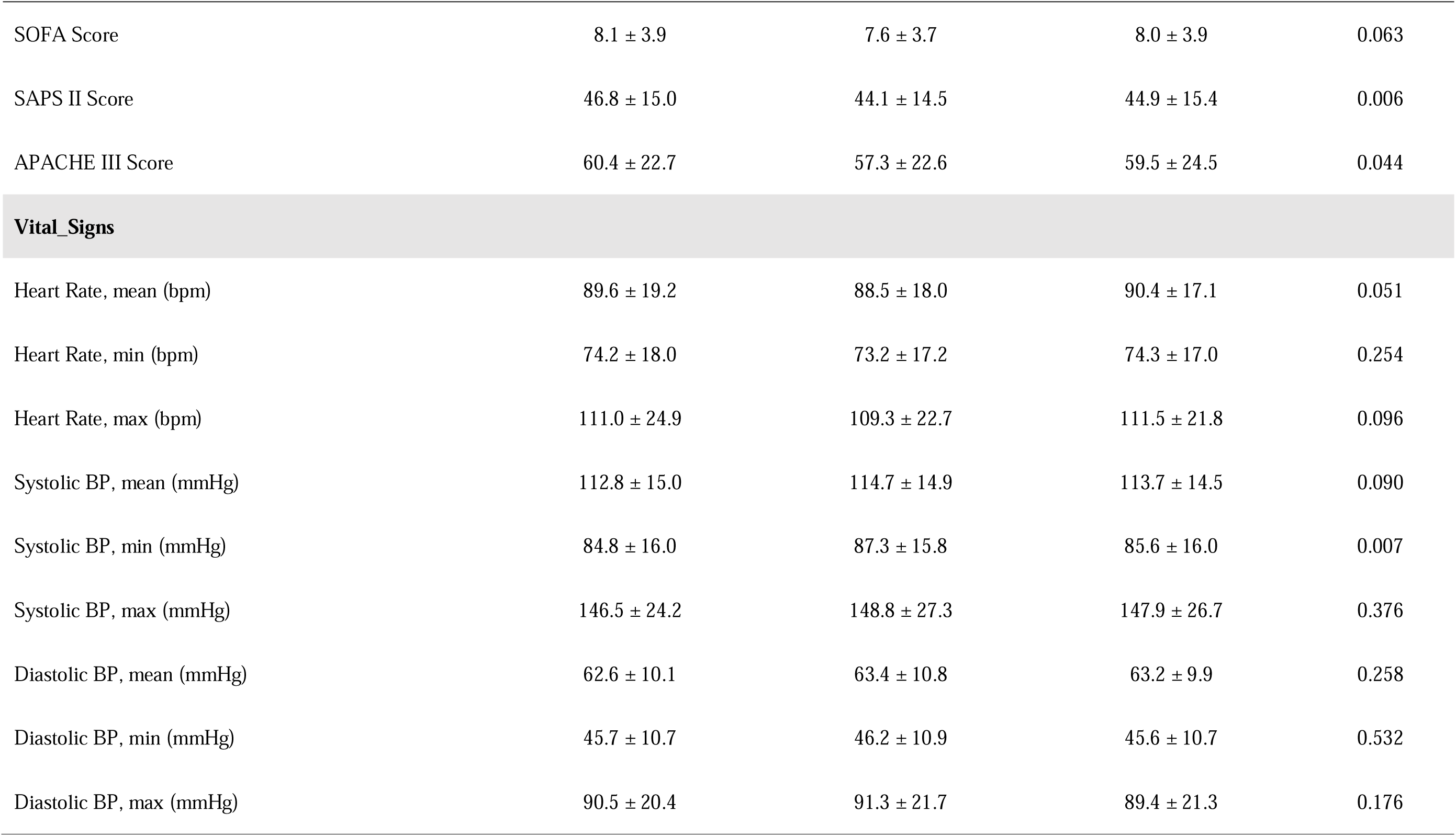
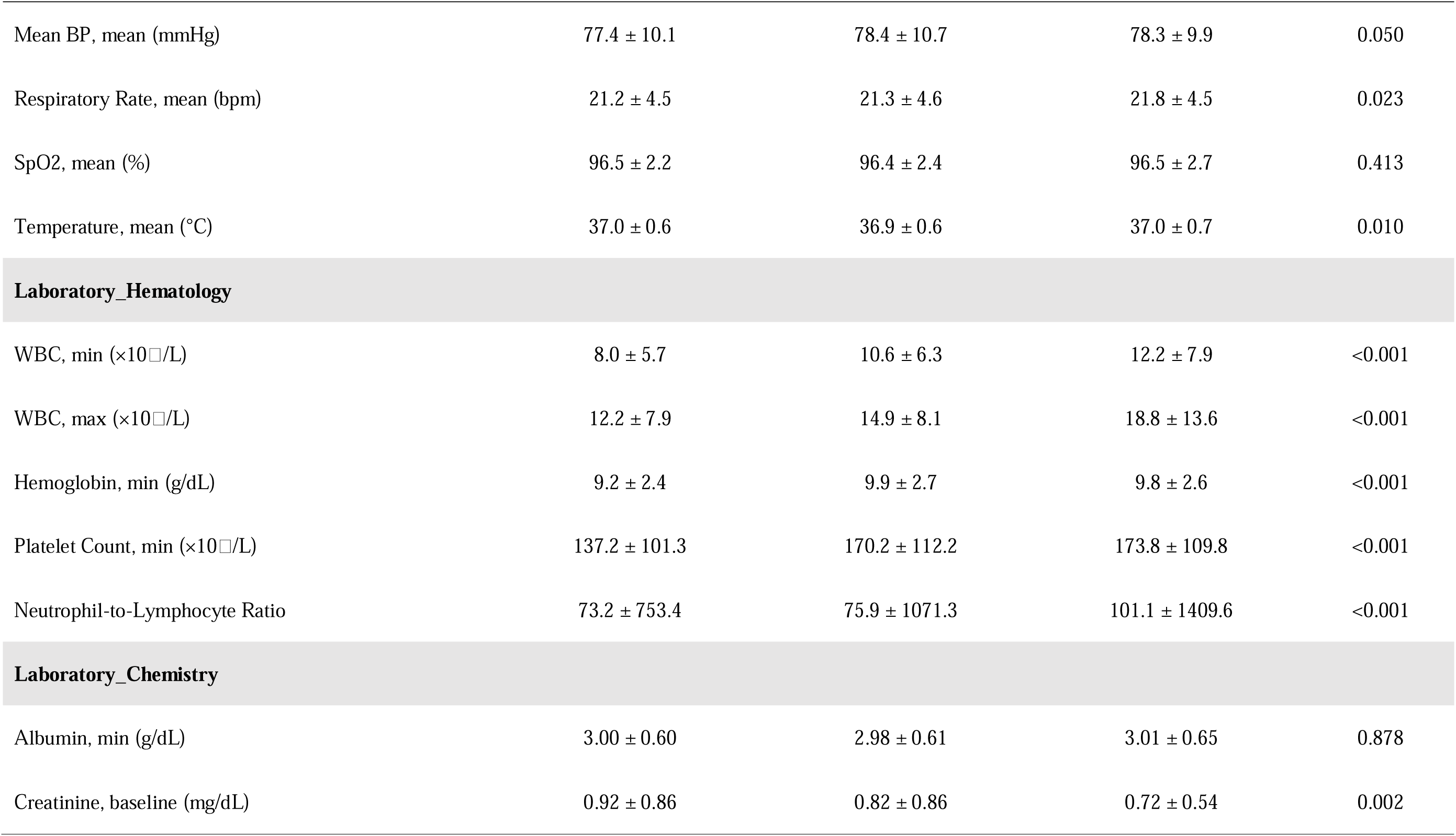
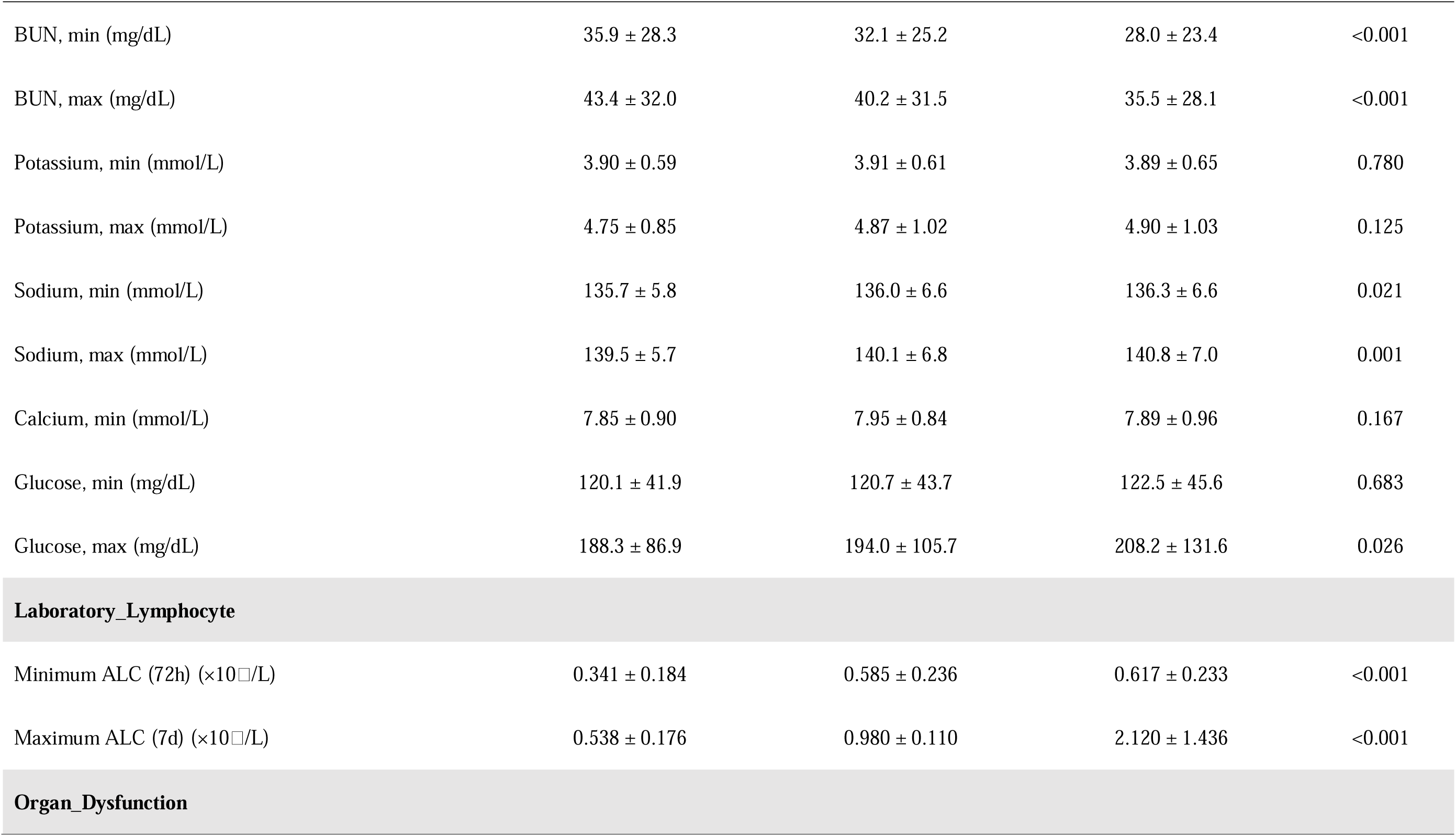
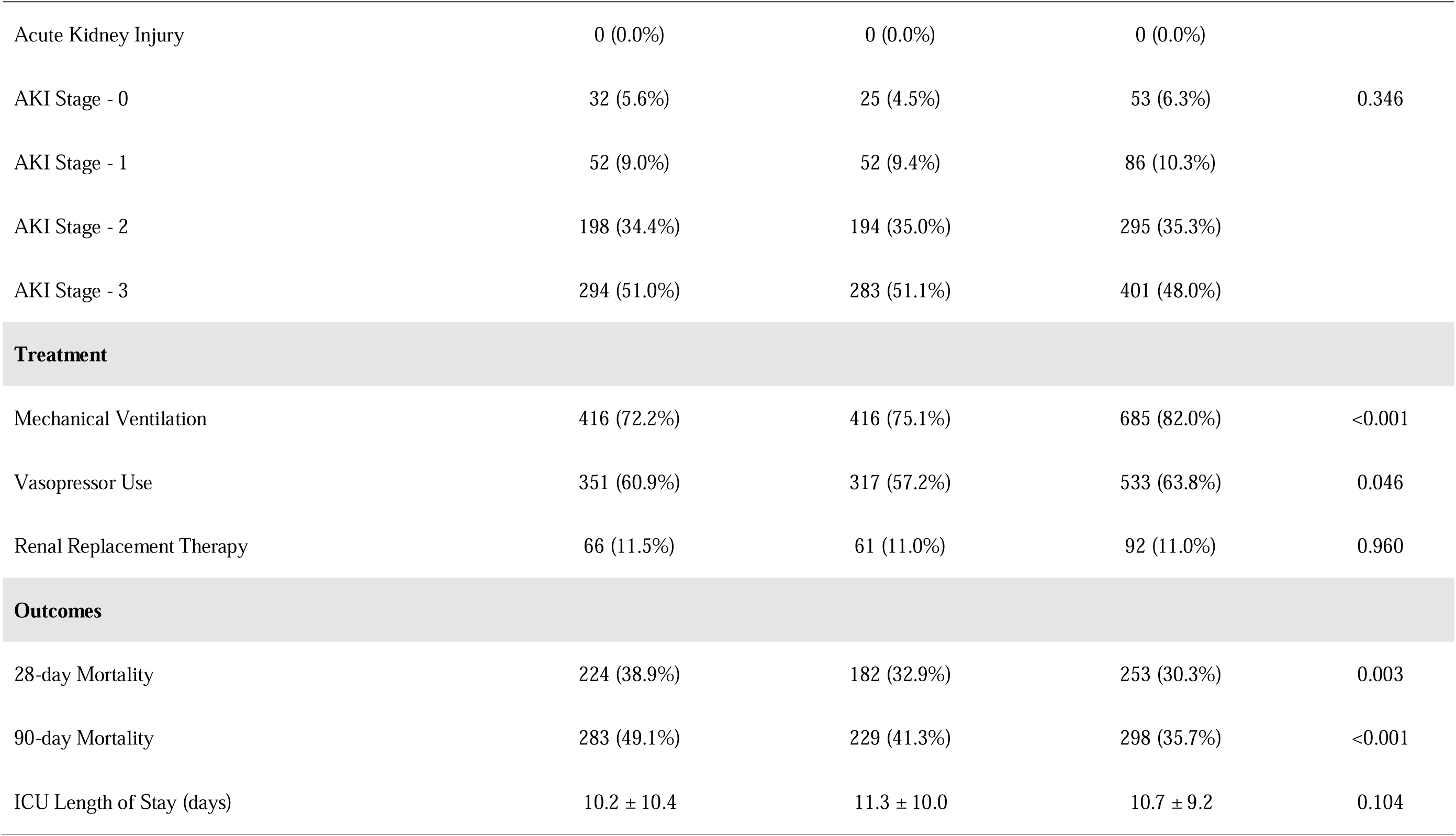
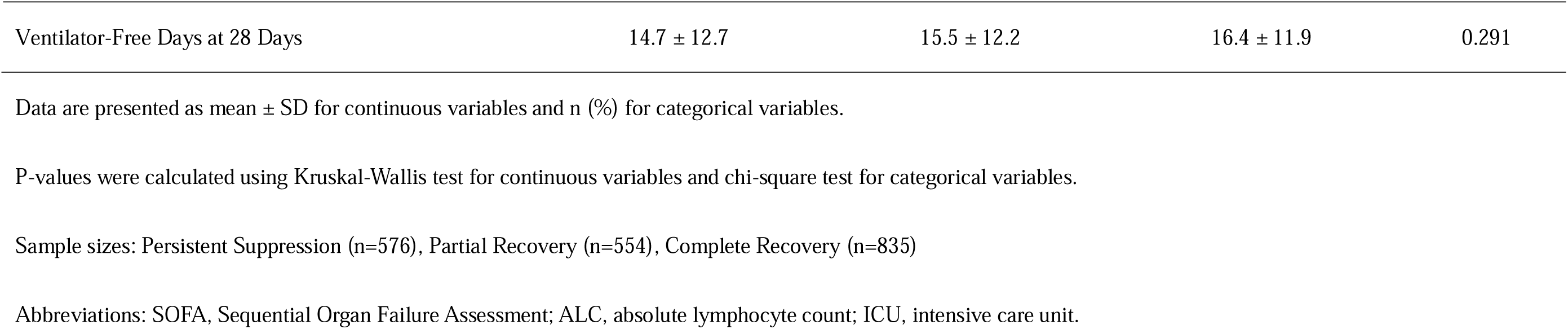
Baseline Characteristics by Lymphocyte Recovery Pattern.

### Association of Lymphocyte Recovery Patterns with 90-Day Mortality

The primary outcome of 90-day all-cause mortality varied significantly across the three groups. The mortality rate was highest in the Persistent Suppression group at 49.1%, followed by the Partial Recovery group at 41.3%, and was lowest in the Complete Recovery group at 35.7% (p < 0.001).

The Kaplan-Meier survival curves visually demonstrate this graded risk, showing that patients in the Persistent Suppression group had a significantly lower 90-day survival probability compared to the other two groups (Log-rank p < 0.001) (Figure 2).

**Figure 2.**
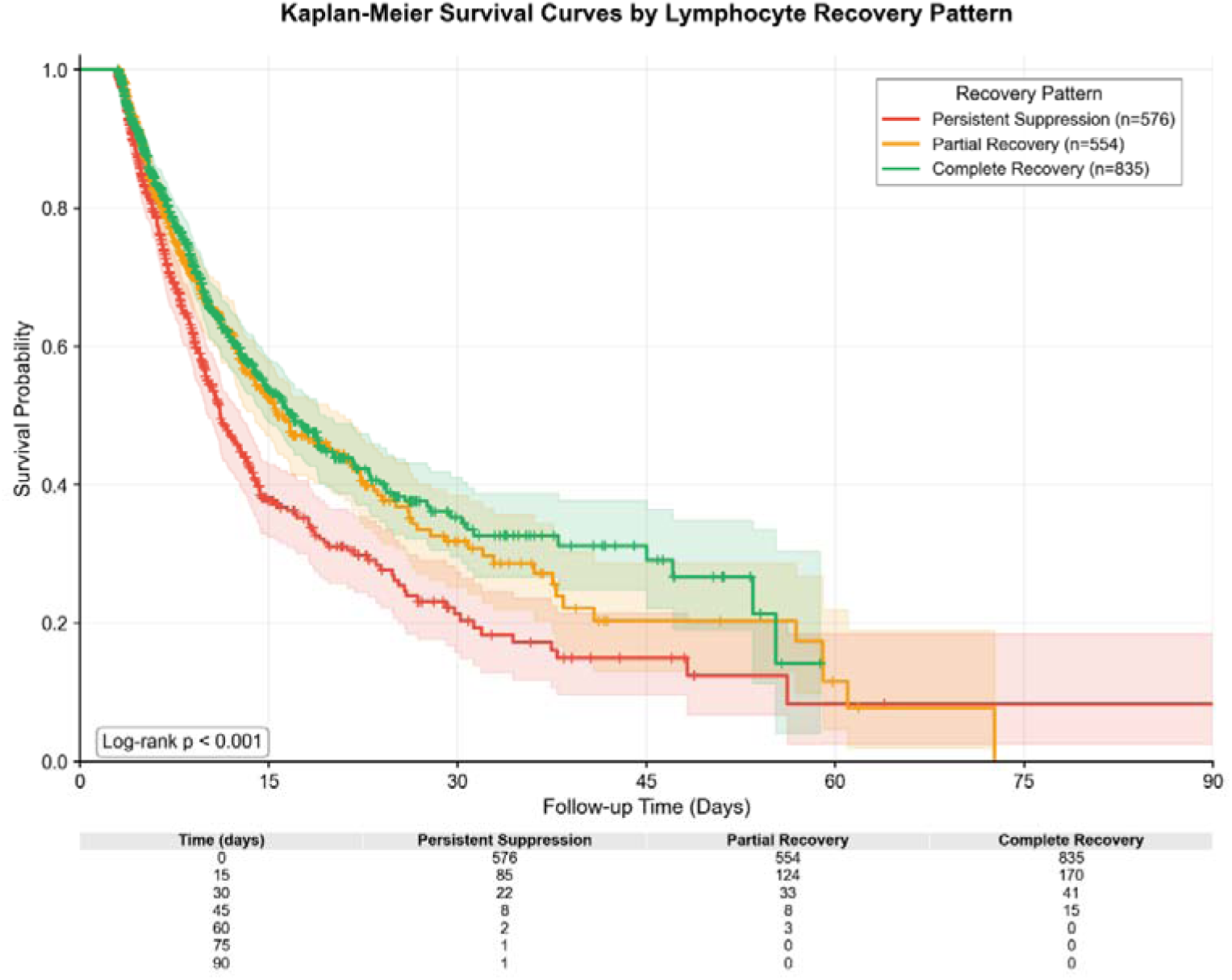
Kaplan-Meier Survival Curves by Lymphocyte Recovery Pattern.

To quantify this association while adjusting for confounders, we constructed multivariable Cox proportional hazards models (Table 2). After adjusting for age, sex, race, Charlson Comorbidity Index, SOFA score, mechanical ventilation, vasopressor use, albumin, platelet count, and creatinine (Model 4), the Persistent Suppression pattern remained a strong and independent predictor of 90-day mortality when compared to the Complete Recovery group (adjusted Hazard Ratio [aHR] 1.31, 95% CI 1.10-1.55, p = 0.002). In contrast, the adjusted risk for the Partial Recovery group was not statistically significant (aHR 1.09, 95% CI 0.92-1.30, p = 0.330).

**Table 2.** Association Between Lymphocyte Recovery Pattern and 90-Day Mortality: Cox Proportional Hazards Regression Analysis.

| Recovery Pattern | Model 1 |  | Model 2 |  | Model 3 |  | Model 4 |  |
| --- | --- | --- | --- | --- | --- | --- | --- | --- |
|  | HR (95% CI) | P | HR (95% CI) | P | HR (95% CI) | P | HR (95% CI) | P |
| **Complete Recovery** | 1 (Reference) | - | 1 (Reference) | - | 1 (Reference) | - | 1 (Reference) | - |
| **Partial Recovery** | 1.16 (0.97-1.37) | 0.098 | 1.09 (0.91-1.29) | 0.345 | 1.06 (0.89-1.26) | 0.517 | 1.09 (0.92-1.30) | 0.330 |
| **Persistent Suppression** | 1.46 (1.24-1.72) | <0.001 | 1.36 (1.15-1.61) | <0.001 | 1.27 (1.07-1.50) | 0.005 | 1.31 (1.10-1.55) | 0.002 |

Abbreviations: HR, hazard ratio; CI, confidence interval; SOFA, Sequential Organ Failure Assessment.
Model 1: Unadjusted model.
Model 2: Adjusted for age, sex, and race.
Model 3: Model 2 plus Charlson Comorbidity Index.
Model 4: Model 3 plus SOFA score, mechanical ventilation, vasopressor use, albumin, platelet count, and creatinine.
Note: Lactate (40.2% missing) and infection source were excluded due to high missingness or unavailability.
Reference group: Complete Recovery (n=835).
Total sample: N=1,965; Events=810 (41.2%).

### Secondary Outcomes

We analyzed secondary outcomes using a Fine-Gray competing risk model, with death treated as a competing event (Table 3). The analysis revealed a significant association between lymphocyte recovery patterns and ICU length of stay. Specifically, both the Persistent Suppression group (subdistribution HR [sHR] 0.94, 95% CI 0.90-0.97) and the Partial Recovery group (sHR 0.95, 95% CI 0.92-0.98) had a significantly lower probability of being discharged from the ICU compared to the Complete Recovery group, indicating a longer duration of ICU stay.

**Table 3.**
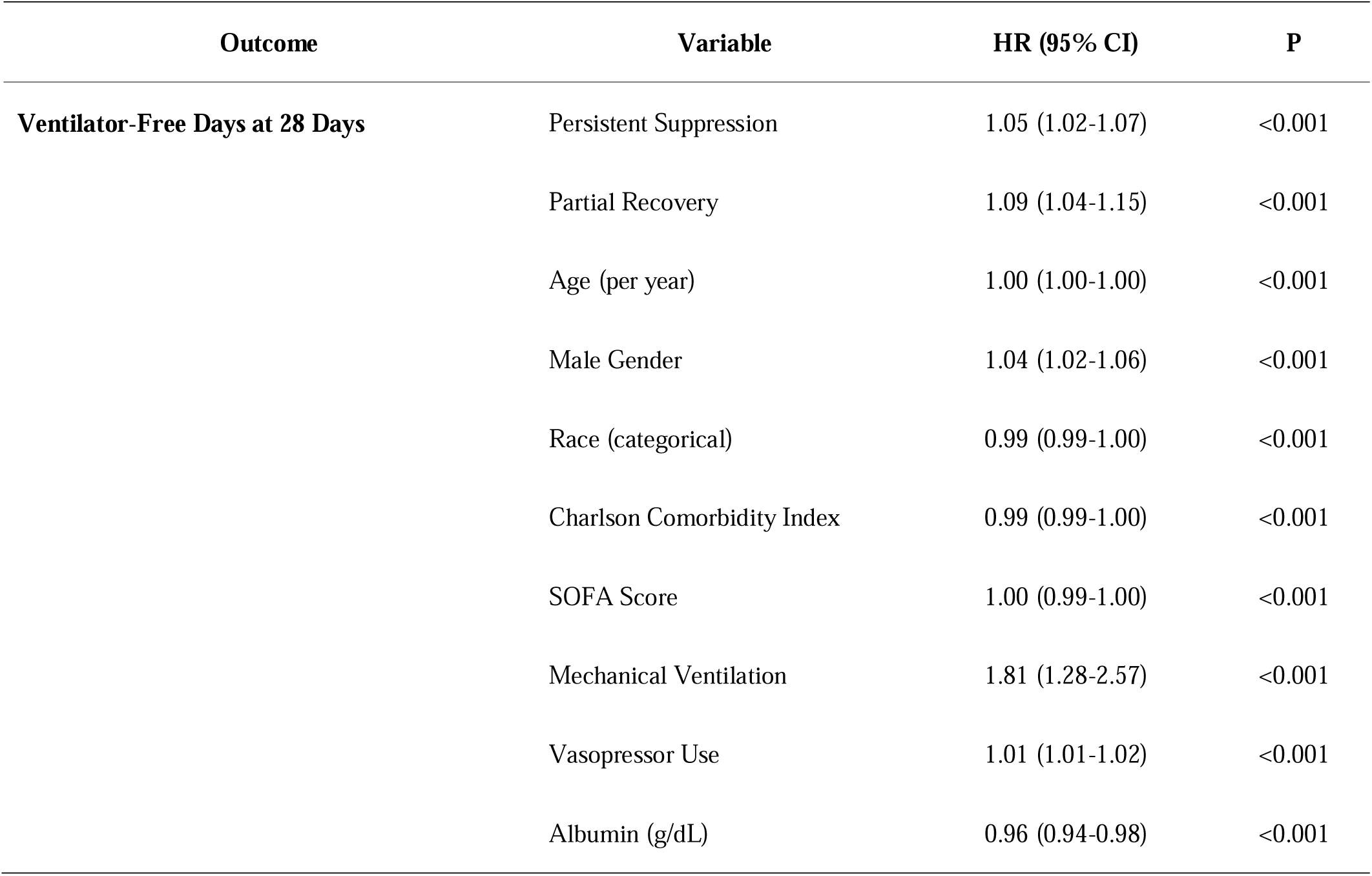
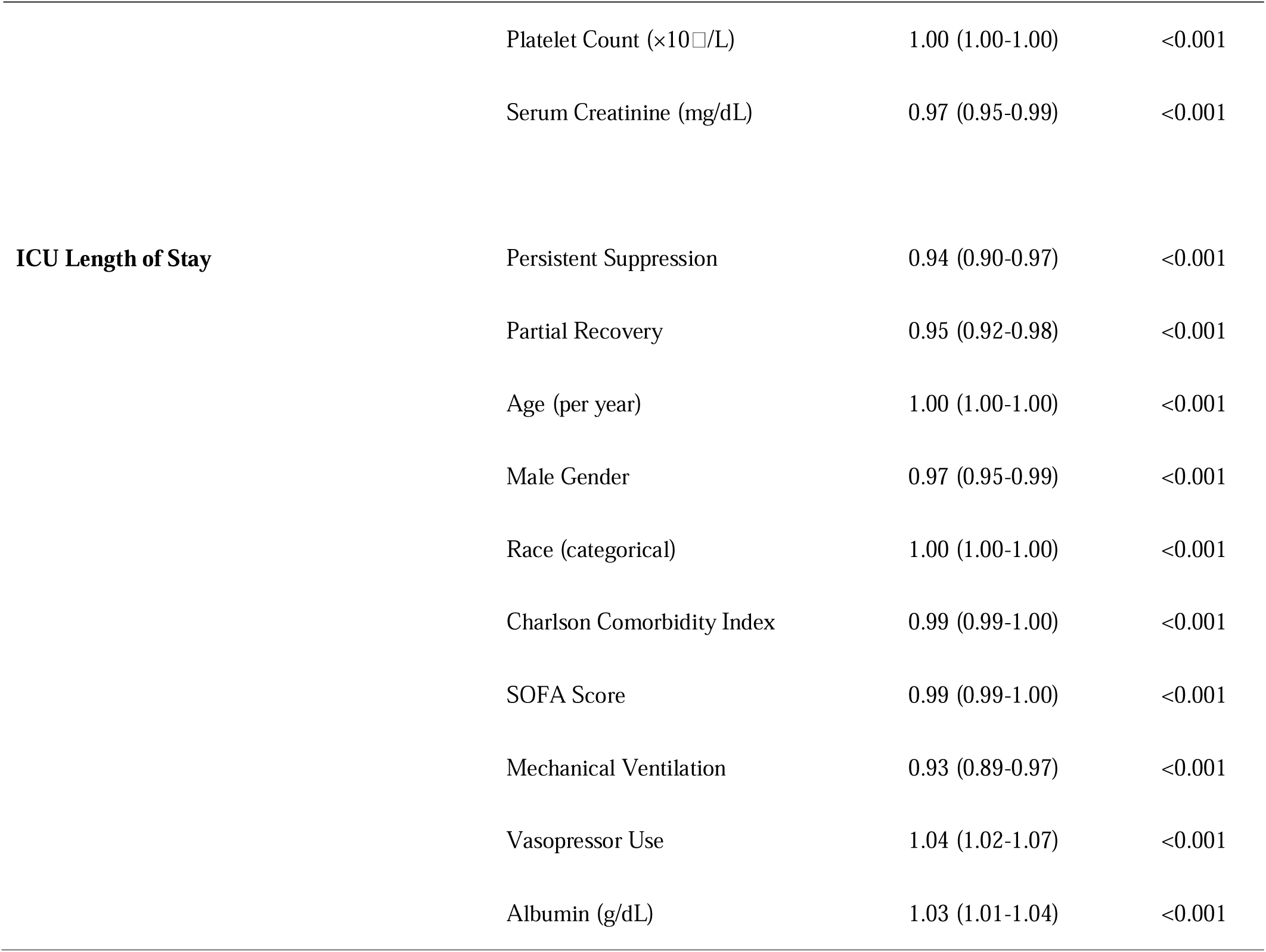
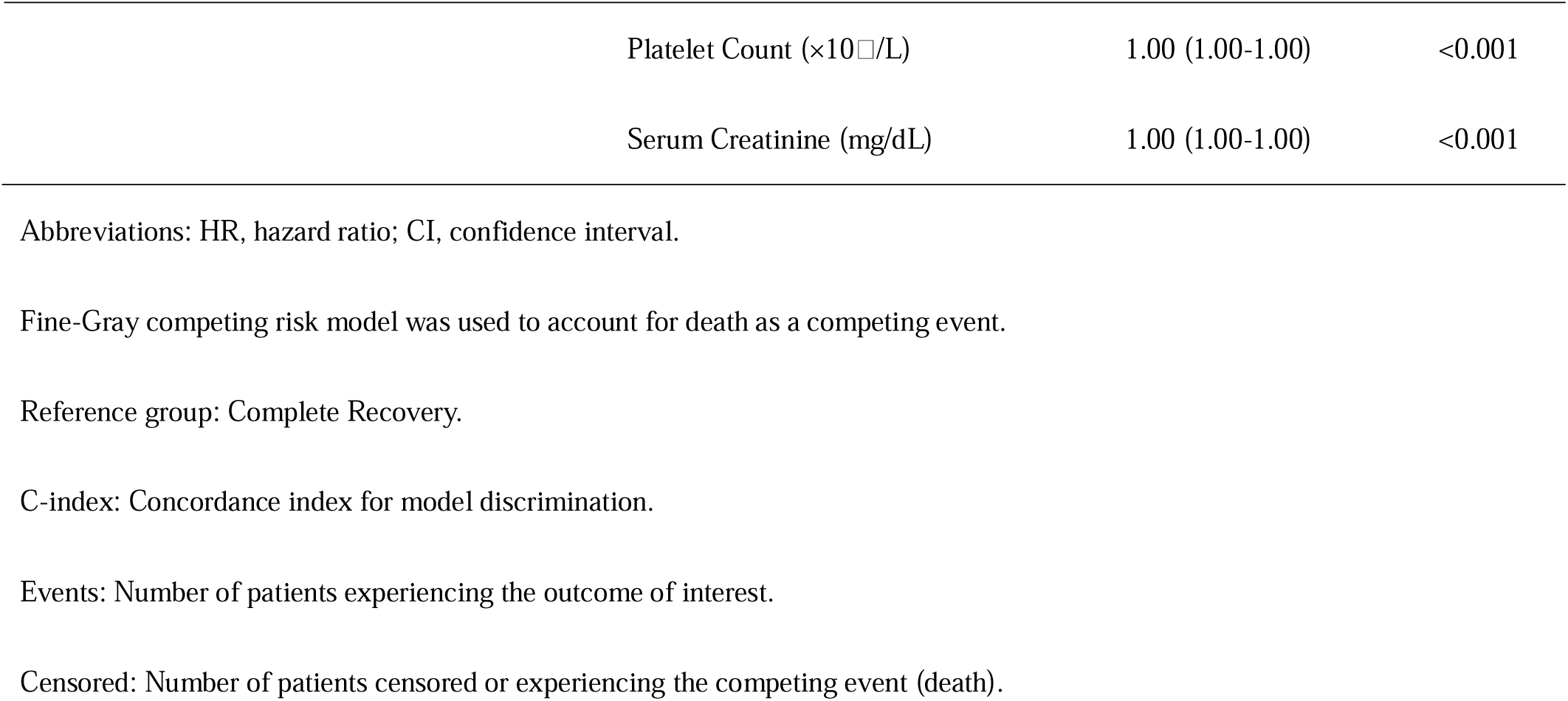
Fine-Gray Competing Risk Model for Secondary Outcomes (Death as Competing Event)

| Outcome | Variable | HR (95% CI) | P |
| --- | --- | --- | --- |
| **Ventilator-Free Days at 28 Days** | Persistent Suppression | 1.05 (1.02-1.07) | <0.001 |
|  | Partial Recovery | 1.09 (1.04-1.15) | <0.001 |
|  | Age (per year) | 1.00 (1.00-1.00) | <0.001 |
|  | Male Gender | 1.04 (1.02-1.06) | <0.001 |
|  | Race (categorical) | 0.99 (0.99-1.00) | <0.001 |
|  | Charlson Comorbidity Index | 0.99 (0.99-1.00) | <0.001 |
|  | SOFA Score | 1.00 (0.99-1.00) | <0.001 |
|  | Mechanical Ventilation | 1.81 (1.28-2.57) | <0.001 |
|  | Vasopressor Use | 1.01 (1.01-1.02) | <0.001 |
|  | Albumin (g/dL) | 0.96 (0.94-0.98) | <0.001 |
| | Platelet Count ($\times 10^9/L$) | 1.00 (1.00-1.00) | <0.001 |
|  | Serum Creatinine (mg/dL) | 0.97 (0.95-0.99) | <0.001 |
| **ICU Length of Stay** | Persistent Suppression | 0.94 (0.90-0.97) | <0.001 |
|  | Partial Recovery | 0.95 (0.92-0.98) | <0.001 |
|  | Age (per year) | 1.00 (1.00-1.00) | <0.001 |
|  | Male Gender | 0.97 (0.95-0.99) | <0.001 |
|  | Race (categorical) | 1.00 (1.00-1.00) | <0.001 |
|  | Charlson Comorbidity Index | 0.99 (0.99-1.00) | <0.001 |
|  | SOFA Score | 0.99 (0.99-1.00) | <0.001 |
|  | Mechanical Ventilation | 0.93 (0.89-0.97) | <0.001 |
|  | Vasopressor Use | 1.04 (1.02-1.07) | <0.001 |
|  | Albumin (g/dL) | 1.03 (1.01-1.04) | <0.001 |

|  |  |  |
| --- | --- | --- |
| Platelet Count ($\times 10^9/L$) | 1.00 (1.00-1.00) | <0.001 |
| Serum Creatinine (mg/dL) | 1.00 (1.00-1.00) | <0.001 |

Abbreviations: HR, hazard ratio; CI, confidence interval.
Fine-Gray competing risk model was used to account for death as a competing event.
Reference group: Complete Recovery.
C-index: Concordance index for model discrimination.
Events: Number of patients experiencing the outcome of interest.
Censored: Number of patients censored or experiencing the competing event (death).

### Predictive Model Performance

An XGBoost machine learning model was developed to predict 90-day mortality, with its performance, interpretability, and robustness detailed in Figure 3.

**Figure 3.**
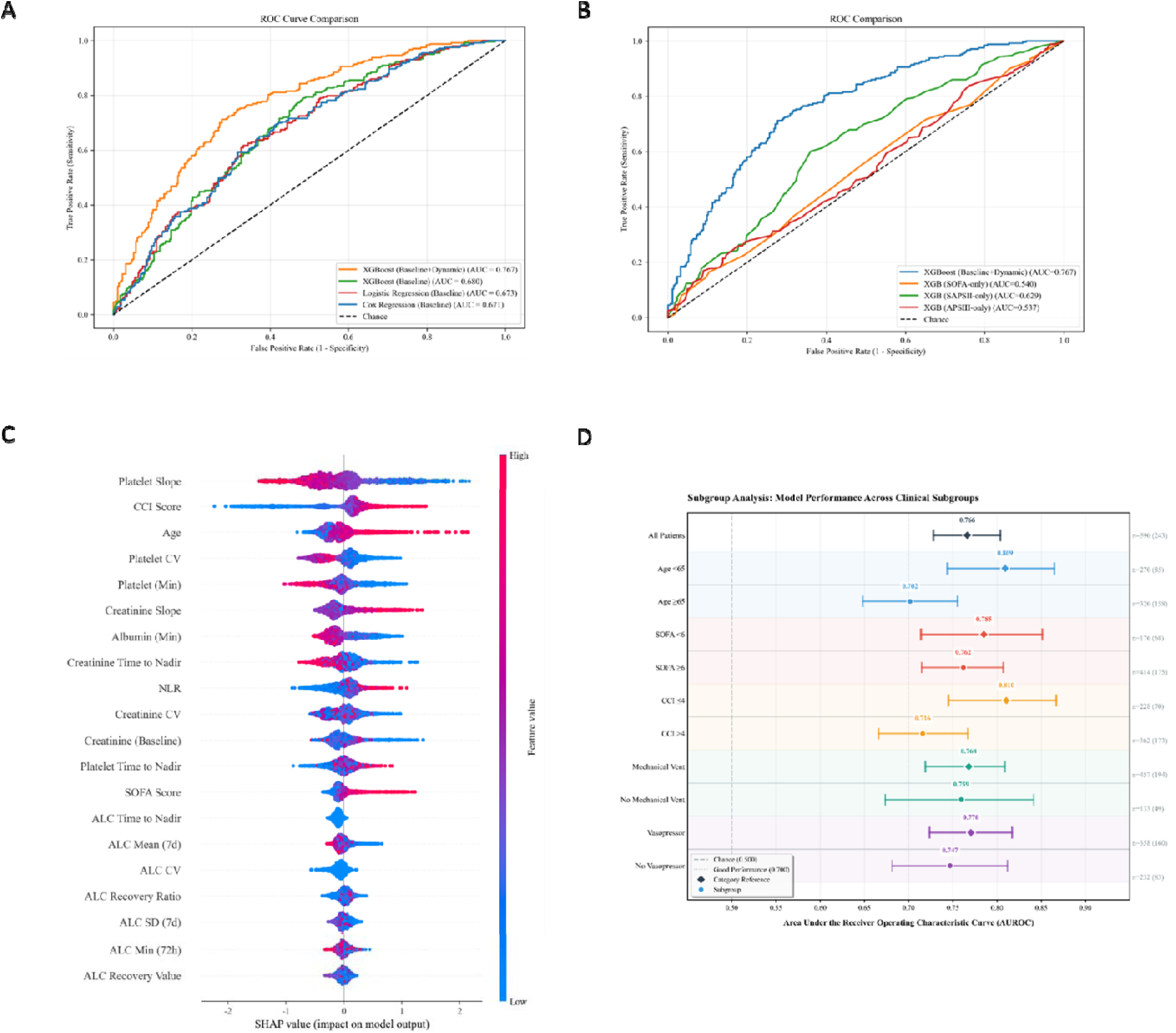
Performance, Interpretability, and Robustness of the XGBoost Model for Predicting 90-Day Mortality. **(A, B)** Comparison of Receiver Operating Characteristic (ROC) curves. The XGBoost (Baseline+Dynamic) model (AUC = 0.767) demonstrated superior discrimination compared to (A) baseline statistical models (e.g., Cox Regression [Baseline], AUC = 0.671) and (B) traditional severity scores (e.g., SOFA-only, AUC = 0.540; SAPSII-only, AUC = 0.629). **(C)** SHAP (SHapley Additive exPlanations) summary plot for global feature importance. The plot ranks the top 20 features contributing to the model’s output. Features are ranked by their mean absolute SHAP value. For each feature, the dot color indicates the feature’s value (red=high, blue=low), and its horizontal position indicates the impact on the model output (higher SHAP value = higher predicted risk of death). **(D)** Forest plot of model performance (AUROC) across predefined clinical subgroups. The model demonstrated robust and good performance (AUROC > 0.700, indicated by the ’Good Performance’ reference line) across most subgroups, including age, SOFA score, and CCI score. Dots represent the AUROC, and horizontal bars represent the 95% CI. **Abbreviations:** ROC, Receiver Operating Characteristic; AUC, Area Under the Curve; XGBoost, Extreme Gradient Boosting; SHAP, SHapley Additive exPlanations; SOFA, Sequential Organ Failure Assessment; SAPS II, Simplified Acute Physiology Score II; CCI, Charlson Comorbidity Index; AUROC, Area Under the Receiver Operating Characteristic Curve.

On the independent test set, the XGBoost model, which integrated both baseline and dynamic clinical features, demonstrated strong discriminative ability with an Area Under the Receiver Operating Characteristic Curve (AUROC) of 0.767 (Figure 3A and 3B). This performance was significantly superior to that of a conventional Cox regression model using only baseline variables (AUROC = 0.671) and far exceeded the predictive capacity of traditional clinical severity scores, such as the SOFA score alone (AUROC = 0.540) or the SAPS II score alone (AUROC = 0.629).

The model showed similarly excellent performance on the training set, achieving an AUROC of 0.851, which also surpassed traditional logistic and Cox regression models (Supplementary Figure 1). Further detailed performance metrics for the test set, including the confusion matrix, precision-recall curve, and calibration curve, are provided in Supplementary Figure 2.

### Model Interpretability and Robustness

To elucidate the XGBoost model’s internal decision-making mechanism, we employed SHAP (SHapley Additive exPlanations) analysisThe SHAP global summary plot (Figure 3C) displays the top 20 features that contributed most to the model’s predictions. The results indicate that Platelet Slope, Charlson Comorbidity Index (CCI Score), and Age were the three most important drivers for predicting 90-day mortality. The core variables of this study—dynamic ALC features such as ALC Mean (7d) and ALC Recovery Ratio—were also identified by the model as significant predictors. This analysis confirms that the model’s decision logic aligns with clinical expectations: lower mean ALC and a poorer recovery ratio (represented by red dots in the plot) consistently pushed the model’s mortality risk prediction higher (SHAP value > 0), demonstrating that the model successfully captured poor ALC recovery as a key risk signal. By analyzing SHAP force plots for three representative patients (Supplementary Figure 3), we can clearly illustrate how the model makes individualized risk decisions: Low-Risk Individual (Supplementary Figure 3A, f(x) = -7.19): This patient’s low-risk prediction was primarily driven by several strong protective factors, including younger age (54.86 years), few comorbidities (CCI Score = 1), and good platelet count recovery (Platelet Slope = 1.51). Given these clear survival advantages, the impact of ALC dynamic features on the final decision was minor.Medium-Low Risk Individual (Supplementary Figure 3B, f(x) = -0.998): This case demonstrates a trade-off between risk and protective factors. Despite a higher comorbidity score (CCI Score = 5), the model ultimately classified the patient as medium-low risk. The key to this decision was the combination of good ALC recovery (ALC Recovery Ratio = 3.61) and a relatively young age (53.87 years), which together acted as the main protective forces, effectively counteracting the influence of other risk factors.High-Risk Individual (Supplementary Figure 3C, f(x) = 5.20): This patient’s extremely high-risk prediction resulted from the accumulation of multiple high-risk features, including a very high comorbidity burden (CCI Score = 11), advanced age (85.21 years), and hypoalbuminemia. On top of these factors, persistent severe lymphopenia (ALC Mean (7d) = 0.3) emerged as the second-largest risk driver, significantly amplifying the model’s final risk assessment.

In summary, the SHAP analysis shows that the dynamic ALC recovery pattern is a critical and clinically interpretable component of the model’s predictive framework. The model not only learned the global rule that "poor ALC recovery increases mortality risk" but also applied this rule precisely at the individual level, treating good ALC recovery as a key protective factor that can mitigate risk and persistent ALC suppression as a key driver that elevates it.

Finally, the model’s robustness was evaluated through subgroup analyses, presented as a forest plot (Figure 3D). The results confirmed that the model maintained good and consistent predictive performance (AUROC > 0.700) across most critical clinical subgroups, including those stratified by age, SOFA score, and Charlson Comorbidity Index.

## Discussion

In this large, retrospective cohort study, we defined three distinct lymphocyte recovery patterns and demonstrated their prognostic significance in patients with sepsis. Our central finding is that a "Persistent Suppression" pattern, characterized by a failure to restore lymphocyte counts, is independently associated with a significantly increased risk of 90-day mortality compared to patients who achieve complete lymphocyte recovery. Furthermore, we developed and validated a machine learning model incorporating these dynamic immunological features, which showed superior predictive performance over traditional clinical scoring systems and static statistical models. These findings offer a new perspective on risk stratification in sepsis, emphasizing the critical value of dynamic monitoring in understanding and predicting patient outcomes. The development of accurate prognostic models is a crucial step toward precision medicine in sepsis, enabling clinicians to identify high-risk patients who may benefit most from targeted interventions, such as immunomodulatory therapies, thereby optimizing resource allocation and improving patient outcomes[18, 19].

Our primary finding—that the dynamic pattern of ALC recovery is an independent predictor of 90-day mortality—provides strong evidence for its utility in monitoring sepsis-induced immunosuppression. It is well-established that a prolonged state of immunosuppression is a key driver of secondary infections and late mortality in sepsis, with lymphopenia being one of its core features[7, 20]. The mechanisms are multifactorial, including extensive apoptosis of T-cells, B-cells, and natural killer cells[21, 22]. While expert consensus already highlights the importance of immune monitoring and recommends ALC as a foundational metric[23], most research has focused on the absolute value or duration of lymphopenia at static time points. Our study innovates by demonstrating that the *capacity for recovery* itself contains superior prognostic information. A state of "Persistent Suppression" likely signifies a failure of the host’s immune system to reconstitute, a condition tightly linked to worse outcomes[24]. This dynamic metric better reflects the functional resilience of the immune system than a single ALC value. Therefore, the ALC recovery pattern can serve as a simple, inexpensive, and dynamic biomarker to identify high-risk patients with sustained immunosuppression. This could be invaluable for patient selection and therapeutic monitoring in future clinical trials of immunomodulatory agents aimed at reversing immune paralysis, such as interleukin-7 (IL-7) or immune checkpoint inhibitors targeting PD-1/PD-L1[6, 25].

Regarding predictive modeling, our XGBoost model demonstrated superior performance (AUROC = 0.767) in predicting 90-day mortality, significantly outperforming widely used standard scores like SOFA (AUROC = 0.540) and SAPS II. Traditional scoring systems primarily rely on physiological variables reflecting acute organ dysfunction, which have limited ability to capture the subsequent state of immunosuppression. In recent years, numerous studies have sought to develop predictive models incorporating novel biomarkers to surpass these traditional scores[26, 27]. The success of our model lies in its effective integration of dynamic immunological features (e.g., platelet slope) with key baseline clinical variables through a machine learning algorithm. The SHAP analysis further revealed that these dynamic features played a crucial role in the model’s decisions, explaining its superior performance compared to models based on static snapshots[28, 29]. Unlike some commercial, poorly performing, or "black-box" predictive models[30], our model is open and interpretable, which is fundamental for clinical trust and application[31]. This tool has the potential to serve as a clinical decision support system, assisting clinicians in identifying patients at high risk for long-term mortality earlier and more accurately[32, 33].

This study has several notable strengths. First, we utilized readily available and inexpensive routine hematological markers (e.g., lymphocyte and platelet counts) to construct our predictors and model. This contrasts with research relying on expensive or technically complex biomarkers (e.g., cytokines, gene expression profiles), making our findings highly practical and translatable to diverse clinical settings[34, 35]. Second, the core innovation of this study is its dynamic perspective. We moved beyond single-point absolute values to analyze data over time, constructing dynamic variables that reflect recovery trends. This aligns with guidance suggesting that the kinetics of biomarkers are more effective than single values for assessing infection severity and treatment response[36]. Our study applies this principle to immune monitoring, proving the superiority of dynamic recovery patterns[37]. Finally, by integrating multiple key clinical features, our prognostic model provides a more comprehensive and robust risk assessment than any single marker alone, embodying a "bench-to-bedside" translational approach[38, 39].

Nevertheless, this study has inherent limitations. First, as a retrospective analysis of an electronic health record (EHR) database, it is susceptible to selection bias and unmeasured confounding; for instance, we could not account for treatments like corticosteroids that may influence lymphocyte counts. A primary limitation of this study is the potential for survivorship bias, stemming from our inclusion criteria which required an ICU stay of more than 72 hours and at least three ALC measurements. This design systematically excludes patients who died early, who may represent the most severe cases of immunosuppression, as well as those who recovered rapidly and were discharged. This selection could lead to an underestimation of the true mortality risk associated with persistent lymphopenia and may affect the generalizability of our model to the entire sepsis population. EHR-based research also faces challenges of data heterogeneity and missing information[40]. Second, our findings originate from a single, large academic medical center’s database. Although the sample size is substantial, the generalizability of our conclusions requires validation in more diverse populations and healthcare settings. The performance of machine learning models often degrades upon external validation; therefore, rigorous testing in multicenter cohorts across different countries and healthcare systems is an essential next step to confirm clinical utility[41, 42]. Future research should focus on prospectively validating the prognostic value of ALC recovery patterns and assessing whether an early warning system based on our model can improve clinical outcomes for patients with sepsis.

In conclusion, the dynamic trajectory of lymphocyte recovery is a critical, independent prognostic factor in patients with sepsis. Failure to restore lymphocyte counts identifies a high-risk population with a poor long-term prognosis. Integrating this dynamic immunological marker into machine learning models enhances predictive performance and represents a promising step towards more personalized risk assessment and management in sepsis. Future prospective studies are needed to validate these findings and to explore whether interventions targeted at patients with poor lymphocyte recovery can improve outcomes.

## Data Availability

The datasets generated from MIMIC are available in the MIMIC repositories

https://mimic.physionet.org/

## Declarations

### Author Contribution

Yuwei Huang, Yuzheng Zhang, and Yongli Gao contributed to the study design and conceptualization. Yuwei Huang and Yuzheng Zhang performed the data extraction and statistical analysis. As independent researchers who did not participate in the primary analysis, Zhilei Fan and Qiong Chen reviewed and validated the data analysis process and assisted with the preparation of figures and tables. Yuwei Huang and Yongli Gao wrote the initial draft of the manuscript. Yongli Gao supervised the entire project. All authors reviewed, revised, and approved the final version of the manuscript.

### Declaration of interests

All authors disclosed no potential conflict of interest.

### Funding

No funding was received for conducting this study.

### Data sharing statement

The datasets generated from MIMIC are available in the MIMIC repositories, https://mimic.physionet.org/

### Disclosure Statement on the Use of AI Tools

We hereby disclose that artificial intelligence (AI) tools were utilized in the preparation of this manuscript. Specifically, the flowchart design and language polishing was assisted by Gemini and Elavax, an AI-based writing assistant, to enhance grammatical accuracy, linguistic fluency, and academic expression consistency.

## Acknowledgements

We thank our colleagues from the Innovation Research Institute and the Academic Department at MedSci for their academic guidance, insightful discussions, and assistance.

## Supplementary Materials

**Supplementary Figure 1.**
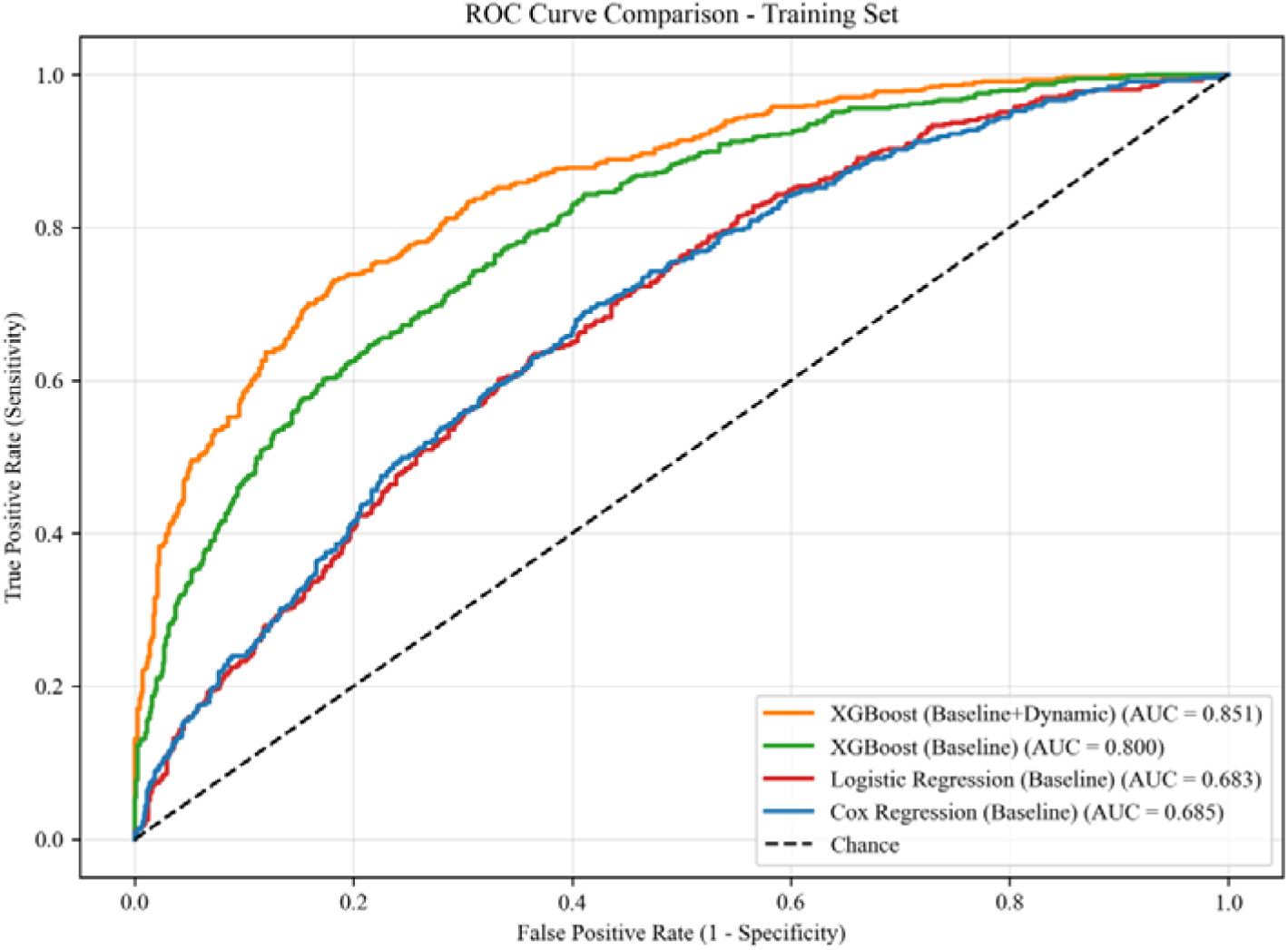
ROC Curve Comparison of Predictive Models in the Training Set.

**Supplementary Figure 2.**
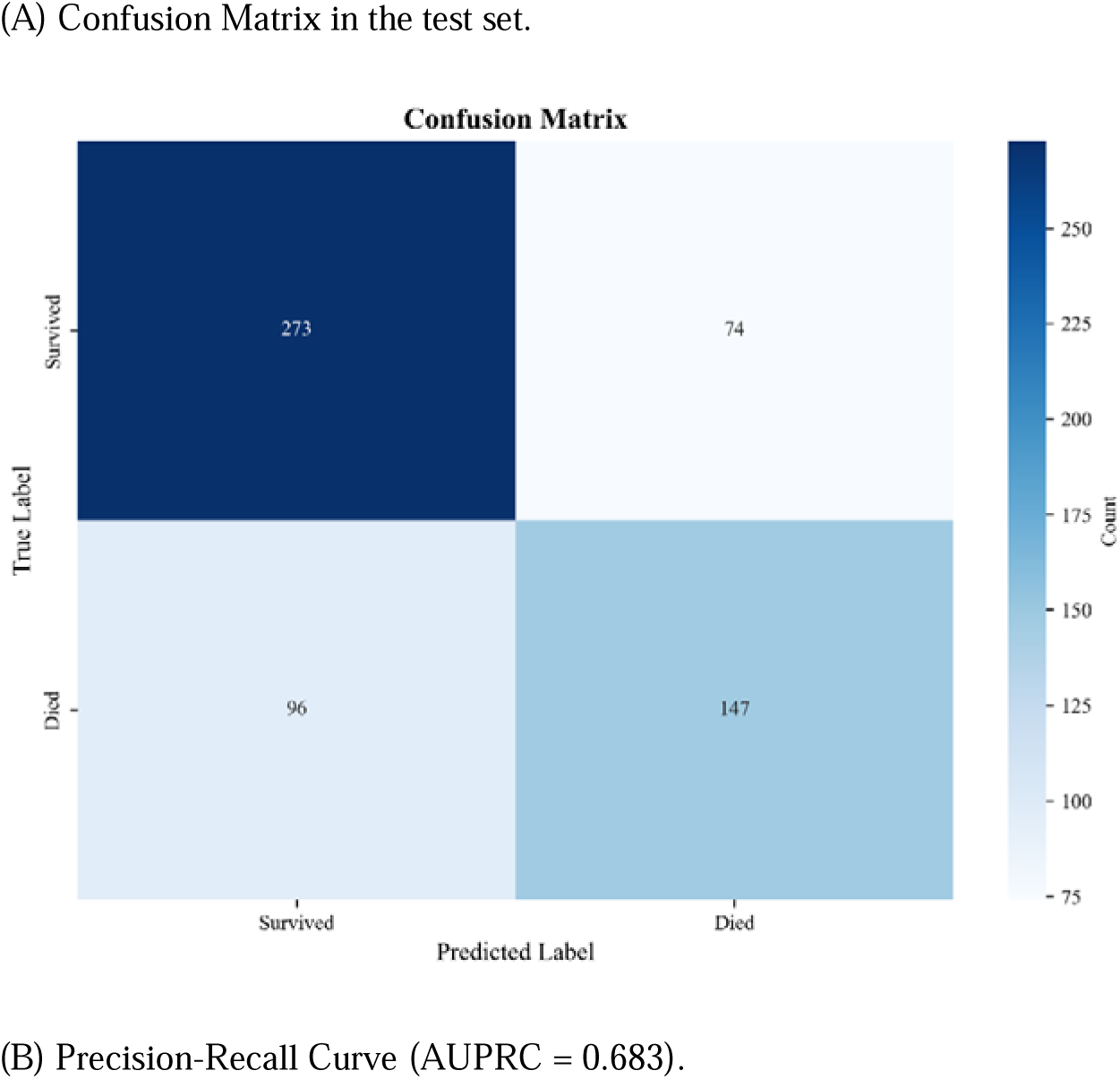
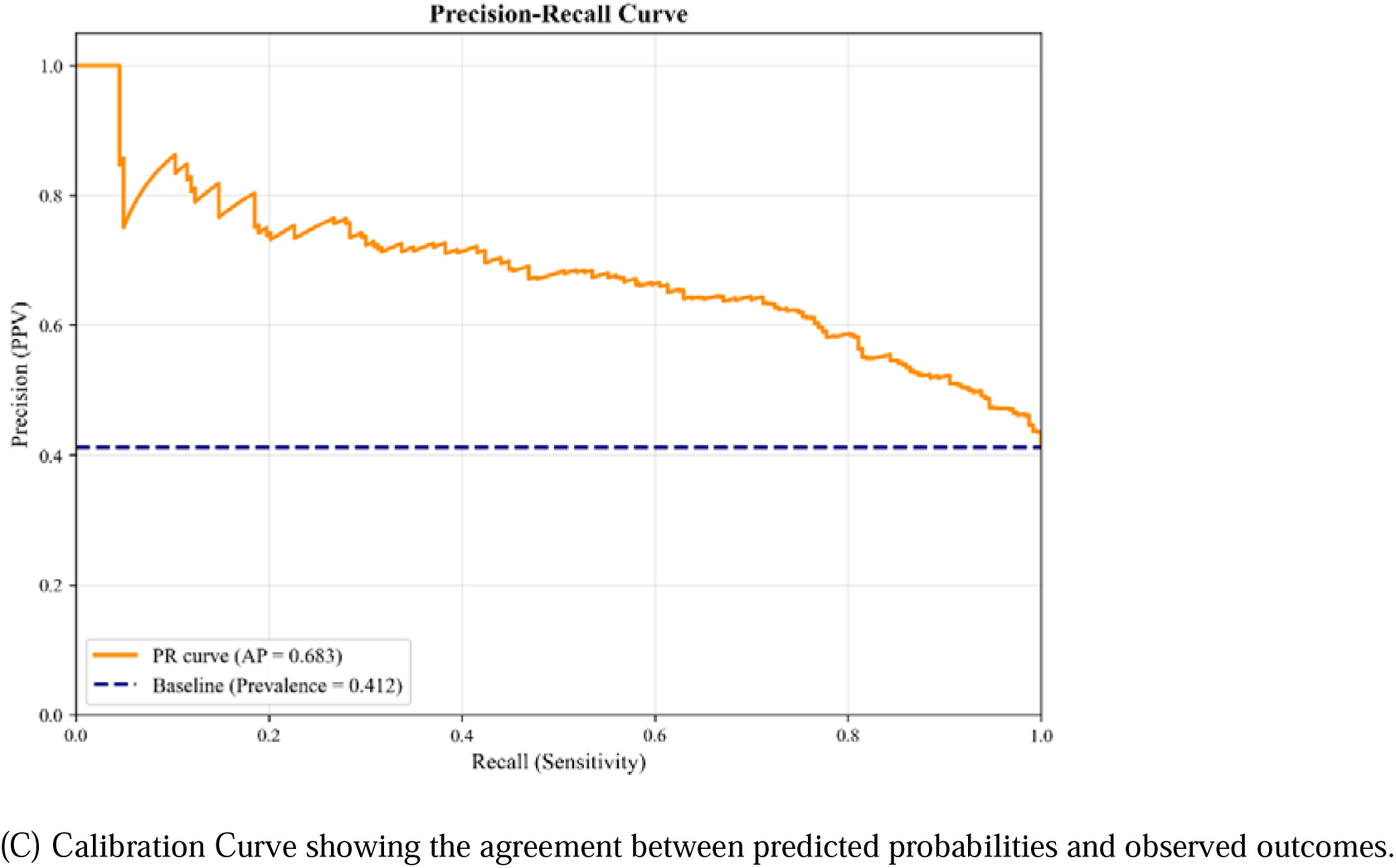
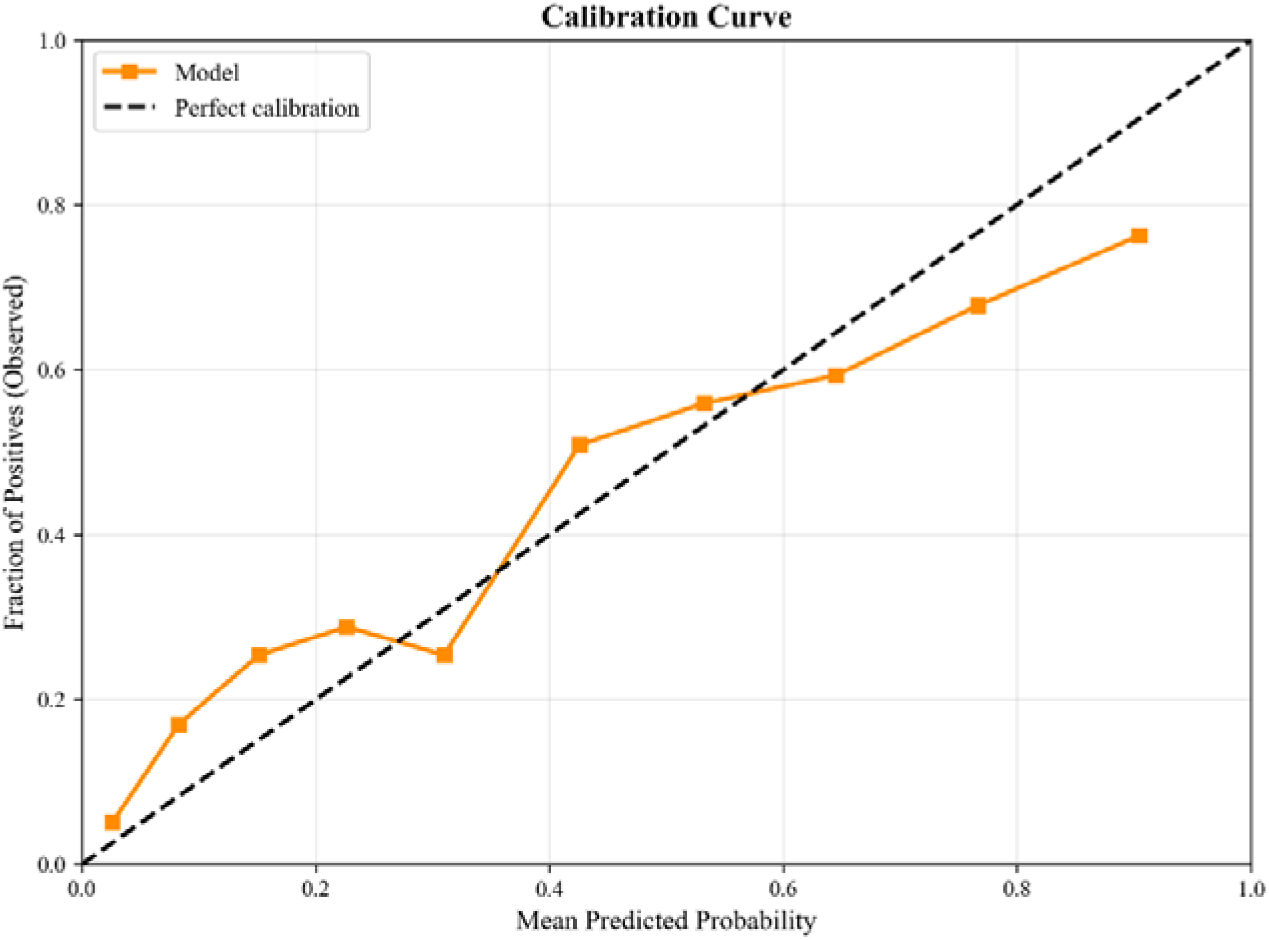
Additional Performance Metrics for the XGBoost Model.

**Supplementary Figure 3.**
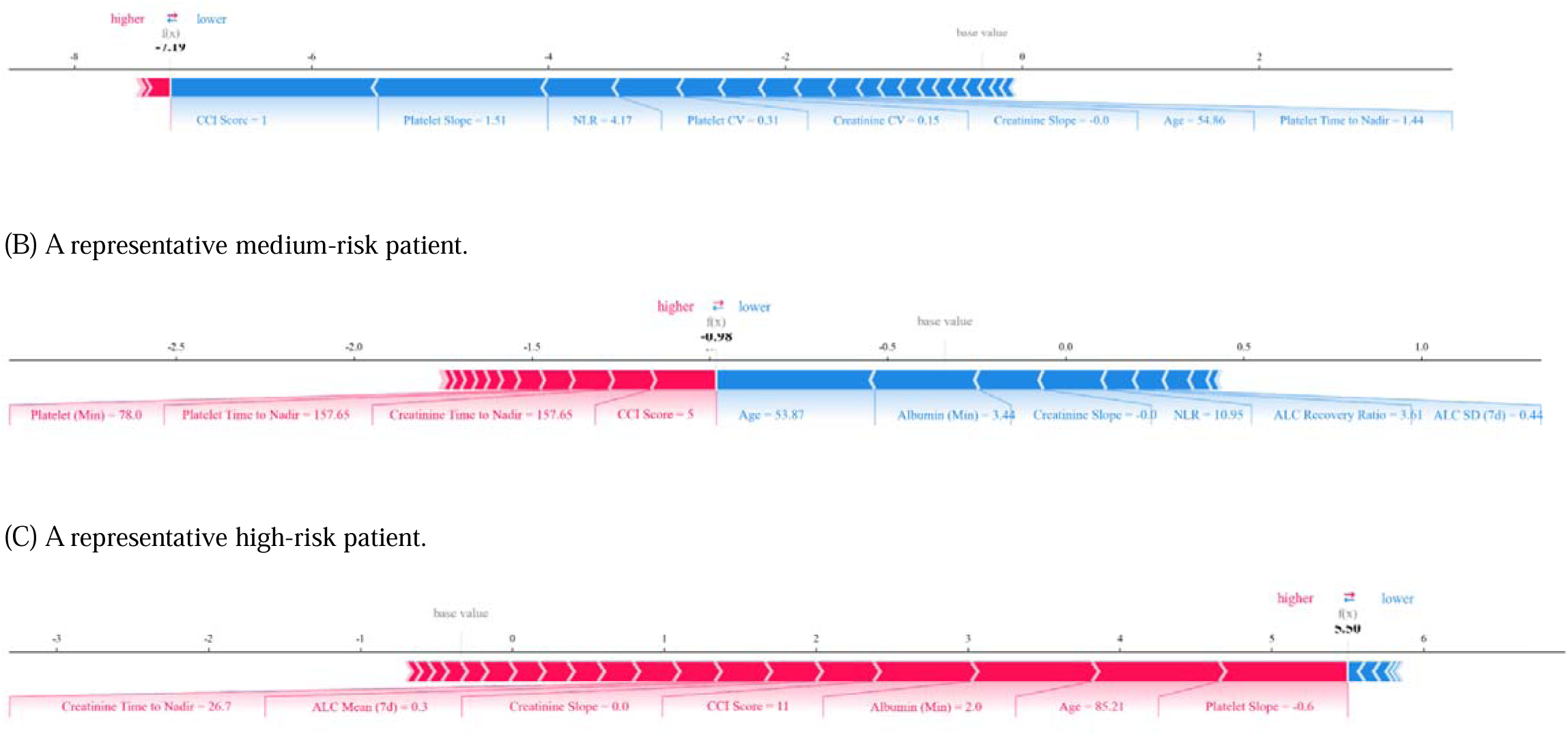
SHAP Force Plots for Individual Patient Explanations. *Legend:* These plots illustrate how different feature values (e.g., CCI Score, Platelet Slope) contribute to (push) the model’s prediction from the base value (average prediction) to the final output (f(x)) for individual cases. Red arrows indicate features increasing risk; blue arrows indicate features decreasing risk.

**Supplementary Table 1.** Detailed Subgroup Analysis of XGBoost Model Performance.

| Category | Subgroup | Sample Size,<br>n (%) | Events,<br>n | AUROC (95%<br>CI) | AUPRC |
| --- | --- | --- | --- | --- | --- |
| Overall | All Patients | 590 (41.2%) | 243 | 0.766<br>(0.728-0.804) | 0.688 |
| Age | Age <65 | 270 (31.5%) | 85 | 0.809<br>(0.744-0.865) | 0.695 |
| Age | Age ≥65 | 320 (49.4%) | 158 | 0.702<br>(0.648-0.755) | 0.693 |
| Disease Severity | SOFA <6 | 176 (38.6%) | 68 | 0.785<br>(0.714-0.851) | 0.679 |
| Disease Severity | SOFA ≥6 | 414 (42.3%) | 175 | 0.762<br>(0.715-0.807) | 0.7 |
| Comorbidity | CCI ≤4 | 228 (30.7%) | 70 | 0.810<br>(0.745-0.867) | 0.701 |
| Comorbidity | CCI >4 | 362 (47.8%) | 173 | 0.716<br>(0.666-0.767) | 0.692 |
| Mechanical<br>Ventilation | Mechanical<br>Vent | 457 (42.5%) | 194 | 0.768<br>(0.719-0.809) | 0.713 |
| Mechanical<br>Ventilation | No Mechanical<br>Vent | 133 (36.8%) | 49 | 0.759<br>(0.673-0.841) | 0.621 |
| Vasopressor Use | Vasopressor | 358 (44.7%) | 160 | 0.770<br>(0.724-0.817) | 0.721 |
| Vasopressor Use | No Vasopressor | 232 (35.8%) | 83 | 0.747<br>(0.681-0.812) | 0.627 |

## Notes

### Competing Interest Statement

The authors have declared no competing interest.

### Funding Statement

This study did not receive any funding.

### Author Declarations

The authors obtained access to the MIMIC-IV database after completing the CITI 'Data or Specimens Only Research' course (Record ID: 66998740) and signing a data use agreement (DUA) with PhysioNet.

